# Presence of ventriculoperitoneal and lumbar shunts stimulate long lasting non-inflammatory changes in the cerebrospinal fluid distinct from the response to bacterial infection

**DOI:** 10.1101/2022.10.12.22280998

**Authors:** Simone M. Cuff, Joseph P. Merola, Matthias Eberl, William P. Gray

## Abstract

Ventriculoperitoneal (VP) shunts are effective at relieving hydrocephalus but are prone to malfunction. There are two hypotheses as to how shunts may malfunction independently of mechanical failure or blockage by debris from initial placement. The first is that the presence of a foreign object results in cells migrating into and colonising the shunt. The second is that the shunts contain either small numbers of live bacteria or residual bacterial products from manufacture or handling, triggering an inflammatory response that attracts cells to the site which go on to cause malfunctions. The presence of bacteria can be difficult to definitively rule in or out, given that they are capable of forming biofilms which poses challenges for isolation and microbiological culture. In this study, we measured 91 soluble immunological molecules and 91 soluble neurological molecules in CSF of patients with VP shunts and compared them to both patients without shunts and those with bacterial infection to determine whether there is an ongoing inflammatory response to shunting. We find that shunts elicit a soluble signature of neural wound healing and cell migration proteins that is distinct from the inflammatory signature of patients with neurological infection. This appears to represent a long-term response, persisting for at least 5 years in one patient.

## Introduction

In the United Kingdom (UK), approximately 3000 shunt operations are carried out each year for a variety of cerebrospinal fluid (CSF)-related disorders. The majority of proximal catheters are placed into the lateral ventricles (76.5%), and the most common indication for revision surgery is infection, especially within the first year (Shunt Registry, UK 2017). Shunt blockage occurs in up to 1 in 20 patients without clear evidence of infection ^5^ and between 1 in 20 and 1 in 8 patients in whom there is clinical evidence of bacterial infection ^5–7^.

The insertion of a shunt catheter is invasive to the central nervous system and results in a localised inflammatory response. Previous studies have demonstrated colonisation of shunt catheters by neuroimmune cells and ingress by astrocytes and glial cells contributing to shunt blockage^1-4^.

It is hypothesised that the stimulus for the cells colonising the shunt surface is either a spontaneous response of astroglia and microglia cells to the presence of a foreign body, or small numbers of bacteria creating biofilms. Biofilms form when bacteria grow on a surface and differ in their growth and gene expression from free-living bacteria ^8–10^. *Staphylococcus*, which is one of the best studied biofilm-forming bacteria and also one of the more common shunt contaminants, exudes polysaccharides as the biofilm begins to form, which help adhere the bacteria to the site while simultaneously rendering the bacteria more resistant to phagocytosis by immune cells such as neutrophils and macrophages ^11,12^.

Shunt infection is difficult to diagnose against a background of significant neurological disease and an inflammatory response to surgery. A recent, large randomised trial reported a 6% rate of shunt infection in 1594 patients (Mallucci et al, 2019). However, biofilm-forming bacteria colonising shunt catheters pose a significant challenge since patients often present with an indolent clinical picture. The low-grade inflammatory response and low bacterial cell count render gold standard bacteriological culture futile producing false negative results. However, they may be detectable indirectly by measuring the resultant inflammatory immune response ^13^.

In this study, we measured a wide range of immunologically and neurologically relevant proteins in the CSF in response to the presence of shunts in patients with no sign of infection and patients with bacterial infection. These were compared to the levels present in the CSF of individuals without shunts. This allowed us to define the nature of the response of the local environment to the presence of shunts and determine whether they are likely to be associated with the presence of biofilms, or due to other causes. Our results suggest that the presence of shunts lead to an increase in biomarkers typical of a non-inflammatory neural wound-healing type response, which was clearly delineated from the inflammation associated with the presence of bacteria.

## Methods

### Patients

Between March 2017 and March 2018, 20 patients in whom CSF sampling was clinically indicated were consented for additional donation along with a matched serum sample for the purpose of this study. Samples were collected and archived via the Wales Neurosciences Research Tissue Bank (WNTRB Ethics Rec Ref: WA/19/0058;Requisition No. 019). CSF samples from consented patients were collected by trained individuals following standardised protocols for aseptic technique. Samples were transferred on ice and then centrifuged (4500rpm, 10min, 4°C) within 30 minutes of collection. The acellular fraction was then stored in 300ul aliquots at -80°C in the WNTRB until analysis.

Patients were categorized into three main groups. Patients with a medically managed neurological condition and no prior shunt history were recruited to group 1 (“No shunt”). This group consisted of 10 patients, all of whom were under regular review for medically-managed idiopathic intracranial hypertension (IIH). The clinical indication for sampling CSF in these patients was therapeutic, i.e. to relieve patients of their symptoms (headache). 3/10 patients subsequently went on to have a VP shunt placed months later.

Patients who had previously undergone a shunt insertion and in whom CSF sampling was clinically indicated for non-infective reasons were recruited to group 2 (“Non-infected Shunted”). This group consisted of 6 patients all with a prior diagnosis of IIH in whom a ventriculoperitoneal shunt was present *in situ*. In 2 of these patients, CSF sampling aided in the diagnosis of a malfunctioning shunt while in the remaining 4 patients it excluded shunt malfunction as the cause for headache.

Four patients were diagnosed with neurological infection and recruited to group 3 (“infected”). Diagnosis was on the basis of a positive Gram stain or, in the case of a negative Gram stain, on the basis of clinical signs consistent with the Infectious Disease Society of America (IDSA) guidelines including fever, meningism and altered consciousness, supported by CSF pleocytosis^14^. In this group, one patient had IIH and the shunt was removed following diagnosis of infection. CSF was sampled from the ventricular catheter on removal, which is the only ventricular sample available from the cohort. Another patient had hydrocephalus secondary to subarachnoid haemorrhage, the remaining two suffered pseudomeningoceles following posterior fossa surgery (one haemangioblastoma and one metastatic tumour).

Linked-anonymised clinical data were collected and stored for subsequent analysis. This included patient demographics, past medical history, concurrent medication, diagnoses, use of antibiotics, method of sampling, indication for sampling, serum and CSF cell counts (haemoglobin, white cell count, red cell count), and the results of extended microbiological culture. Patient characteristics are described in **Table 1**.

**Table 1.** Patient characteristics. Data represented as categorical data or as mean ± SD (range).*= insufficient cells for meaningful % polymorphonuclear granulocytes to be determined.

|  | No shunt<br>(n=10) | Non-infected shunt<br>(n=6) | Infection<br>(n=4) |
| --- | --- | --- | --- |
| Gender (M/F) | 0/10 | 1/5 | 2/2 |
| Age at inclusion (years) | 33.8 ± 7.2 years | 34.3 ± 9.3 years | 43.0 ± 3.7 years |
| Comorbidities (y/n) |  |  |  |
| Diabetes | 1/10 | 0/6 | 0/4 |
| Hypertension | 0/10 | 0/6 | 1/3 |
| Asthma | 0/10 | 1/5 | 0/4 |
| Fibromyalgia | 1/10 | 1/5 | 0/4 |
| Indication for CSF Sampling | 10 symptom control | 5 symptom control<br>1 revision of shunt | 4 diagnosis of infection |
| Method of CSF Sampling | 10 LP | 5 LP<br>1 shunt revision | 1 LP<br>2 LD<br>1 shunt revision |
| SERUM |  |  |  |
| Haemoglobin (g/l) | n/a | 138.5 ± 18.8 (113-166) | 192.0 ± 82.9 (108-295) |
| Red Blood Cells (x10<sup>9</sup>/ml) | n/a | 4.6 ± 0.3 (4.3-5.1) | 6.5 ± 2.8 (3.5-9.4) |
| White Blood Cells (x10<sup>6</sup>/ml) | n/a | 9.2 ± 2.6 (5.6-12.6) | 16.8 ± 4.2 (11.1-22.5) |
| Neutrophils (x10<sup>6</sup>/ml) | n/a | 5.8 ± 2.3 (3.1-9.1) | 12.4 ± 2.3 (9.7-16.0) |
| Lymphocytes (x10<sup>6</sup>/ml) | n/a | 1.9 ± 0.5 (1.2-2.6) | 2.9 ± 1.5 (0.7-4.3) |
| CSF |  |  |  |
| Red Cell Count (per µl) | 1762.9 ± 9103.6<br>(0-17558) | 2271 ± 4008<br>(2-11304) | 2903 ± 4698<br>(52-11035) |
| White Cell Count (per µl) | 3.83 ± 19.6 (0-42) | 3.8 ± 5.8 (0-16) | 672 ± 469 (30-1350) |
| % Polymorph count | n/a * | n/a * | 91.7 ± 4.9 (86-95) |
| Gram Stain | No organisms seen | No organisms seen | 2x Gram positive cocci |
| Microbiological culture | No growth on<br>extended cultures | No growth on extended<br>culture | 2x coagulase negative<br>*Staphylococcus* |

### Sample analysis

Samples were analysed by Olink Analysis Service (Olink Proteomics, Uppsala, Sweden) using the proximity extension assay (PEA) with Proseek® Multiplex Inflammation I and Neurology panels **(Suppl. Table 1)**. Data were normalised using standard plasma controls and are expressed as normalized protein expression (NPX). NPX represents semi-quantitative log_2_ units between 0 and 15 which reflect relative protein concentration but do not quantify it, making the assay suitable for comparisons between samples but not a definitive quantification in itself.

### Data Analysis

Data analysis was performed in R version 4.0. Graphing was performed in R, GraphPad Prism 9.0 (GraphPad Software, California USA), and Morpheus (https://software.broadinstitute.org/Morpheus). Invariant markers for removal were defined by graphing standard deviations (SD) of all biomarkers and fitting curves to the results. The minimum point between the main curve and the large number of zero and near-zero readings was taken as the cut-off for invariance (cut-off=0.383). All biomarkers with SD below the cut-off were regarded as invariant and removed from further analysis.

## Results

### Patients

Between March 2017 and March 2018, CSF from 20 patients was collected and analysed according to a standardised protocol. Table 1 summarises the baseline characteristics. 10 (50%) patients had no shunt history, while 6 (30%) had a shunt *in situ* but no signs of infection. 4 (20%) patients had clinical symptoms and signs of CSF infection. Patients were overwhelmingly female, consistent with the predominantly IIH demographic (18/20; 90%), with few co-morbidities. 18 (90%) CSF samples were acquired from the lumbar cistern either by way of lumbar puncture or lumbar drain. 2 (10%) samples were taken directly from the ventricles during shunt removal.

In two patients who were infected, the causative organism was identified by microbiological culture as coagulase negative *Staphylococcus* spp. Although Gram stain and extended culture were negative in the remaining two patients they were clinically diagnosed as infected due to CSF pleocytosis coupled with clinical symptoms and raised inflammatory markers.

### The response to infection is distinct from the response to the presence of shunts

To identify differences in soluble CSF biomarkers between those individuals with and without shunts, a total of 182 neurological and immunological protein markers were screened. 39 of those biomarkers were found not to differ between individuals and so were removed from the subsequent analysis **(Supplementary Table 1)**. The remaining 145 features were subjected to a principal component analysis (PCA). Together, principal components 1 & 2 (PC1 & PC2) accounted for 91.0% of total variation between individuals. 68.8% of variation was encompassed by PC1 and reflected the presence of infection, consistent with the high sensitivity of the immune response to bacterial components **(Figure 1a)**. PC2 encompassed 22.2% of variation and distinguished between shunted and unshunted individuals. Clear biomarker patterns could be identified, with highly correlated markers being differentially expressed between groups **(Figure 1b)**. The contribution of markers to each principal component was visualised with a loadings plot **(Figure 1c)**, in which higher loadings values and closer alignment with the axis indicate biomarkers that contribute most to the individual principal components. Principal component 1 was indicative of variation between patients with or without bacterial infection and showed that patients with known bacterial infection had a profile in which cytokines and chemokines had high loading values. In comparison, PC2, which indicated variation due to the presence of shunts without infection, was typified by a distinct profile of largely neurological proteins. These had relatively lower loadings compared to the response to infection. This showed that the response to the presence of shunts could clearly be distinguished from the biomarker response accompanying bacterial infection.

**Figure 1.**
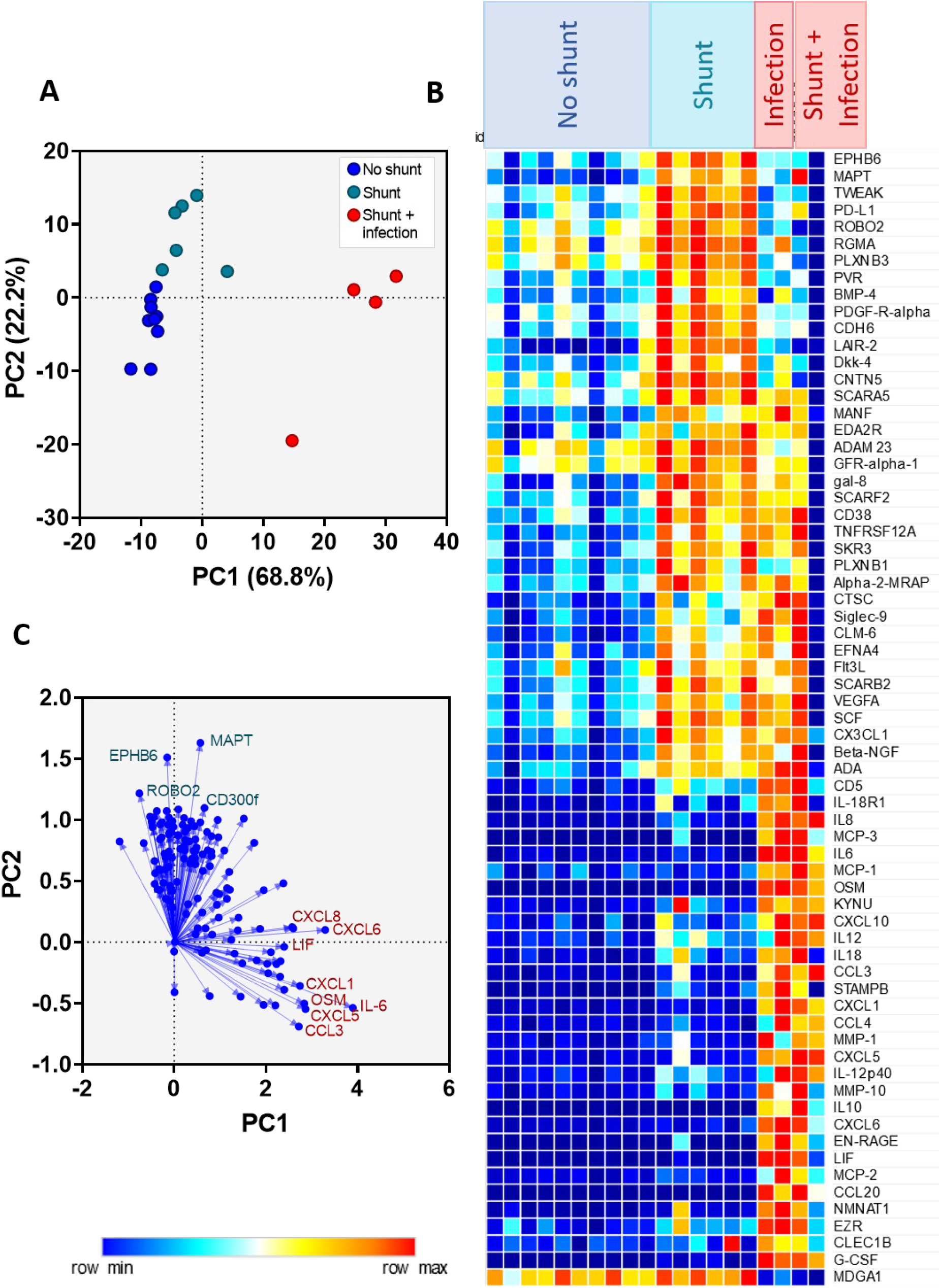
Profile of biomarkers differentiating between shunt status and infection status. (A) PCA separating unshunted patients (dark blue), patients with shunts (teal) and patients with shunts and bacterial infections(red). (B) Heat map displaying biomarkers with contribution of 1 or greater to PC1 or PC2. (C) Loading map showing the contribution of biomarkers to PC1 and 2

### Biomarkers in patients with shunts are associated with cell movement and neural development biomarkers

In order to determine the biological explanation for the changes in biomarker expression, the roles of the proteins were assessed using the Gene Ontology database and critical appraisal of the existing literature. Biomarkers upregulated in shunted individuals were overwhelmingly associated with axonogenesis, neuronal differentiation and cellular locomotion **(Figure 2)**. Interestingly, those proteins associated with axonogenesis and neuronal differentiation were no longer increased in shunted patients where infection was present. The infected individuals instead had uniformly higher levels of immune-associated proteins, as described previously (Cuff et al. 2020). This was consistent with the inflammation seen in the patients. The overarching roles of the proteins is summarised in **Figure 3**, which shows the intersection between the response of the brain to shunts and the immune response to infection.

**Figure 2.**
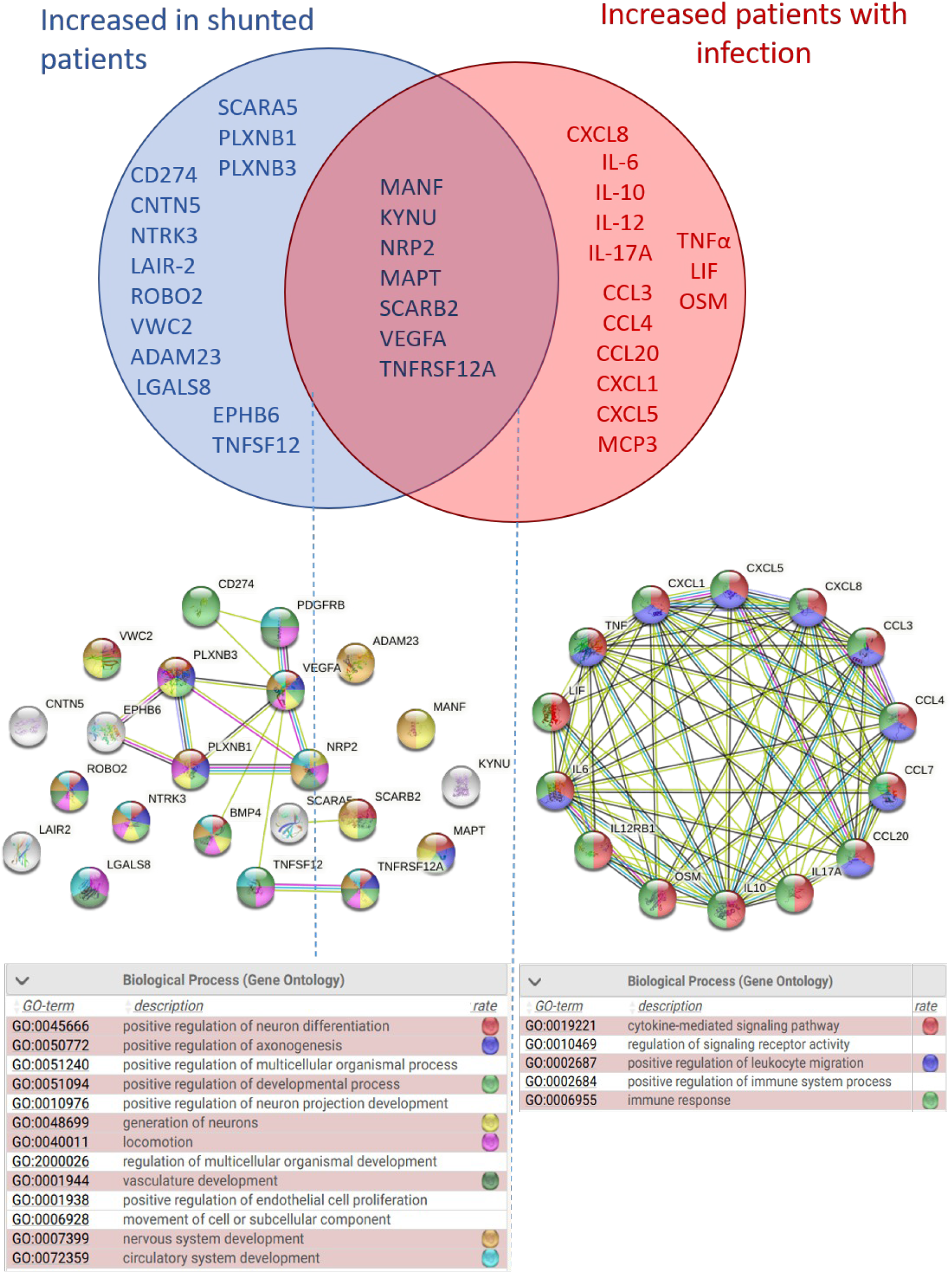
Gene ontology associated with proteins increased in each patient subgroup

**Figure 3.**
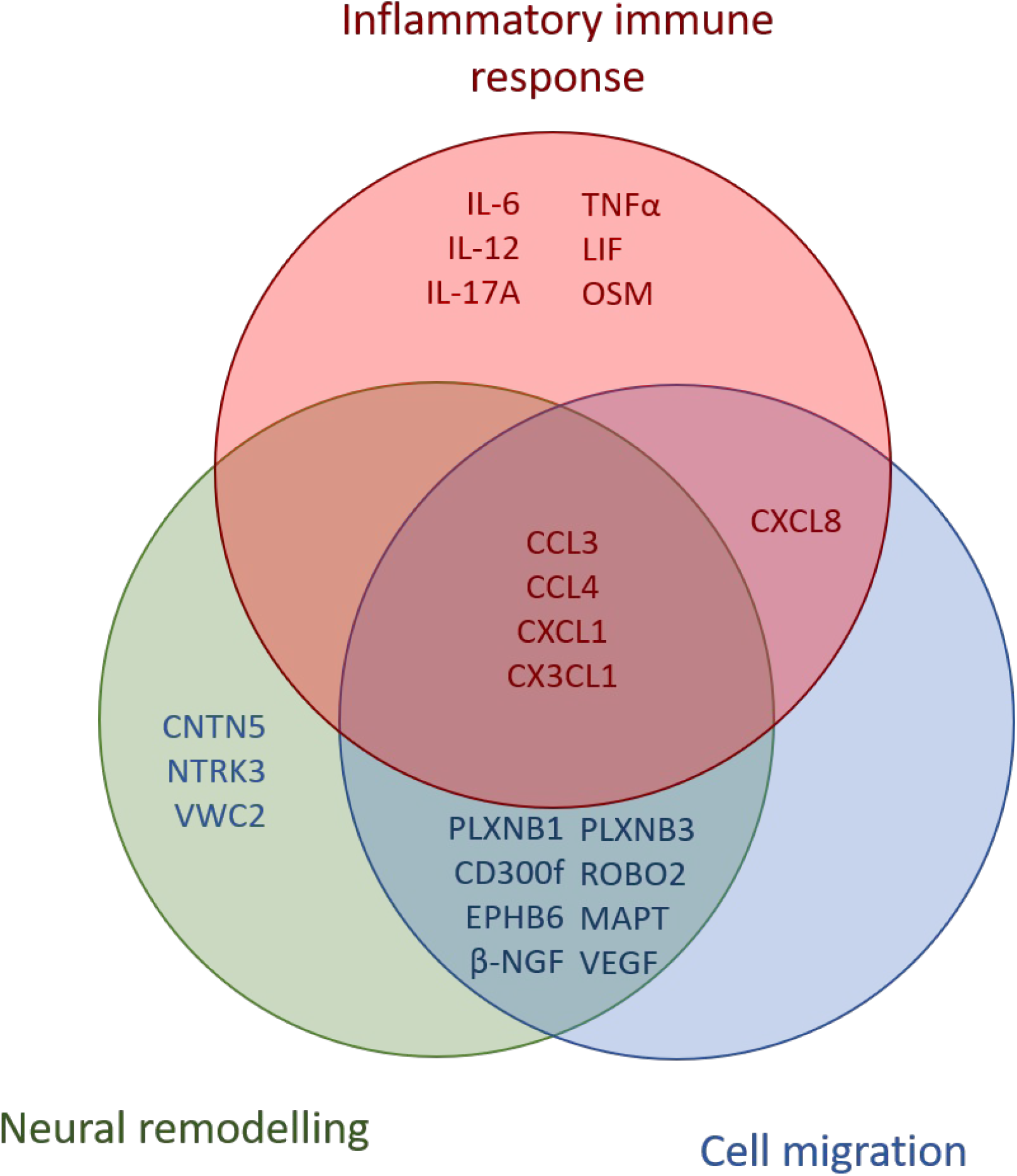
Pathways upregulated in bacterial infection compared to aseptic shunt insertion. Venn diagram showing the most highly implicated pathways of biomarkers upregulated significantly in either bacterially infected (red text) or shunted (blue text) individuals.

### Observed increases in CSF biomarker expression in patients with shunts do not diminish over time

Given that many of the proteins increased in shunted individuals, identified in **Figures 2 and 3**, were important for cell movement and coagulation, we hypothesised that these biomarkers were indicative of a wound healing response to shunt surgery. To test this, CSF biomarker expression was compared according to time since the most recent shunt insertion or revision. Surprisingly, the increase in biomarkers seen in shunted individuals was independent of the time since the insertion or last revision of the shunt. This was despite the time points ranging from less than 1 month to more than 5 years prior to sampling **(Figure 4)**. Thus, we concluded that these biomarkers were not associated with the initial shunt insertion or last revision. Instead, we concluded that the raised biomarkers were indicative of either an acute response to the clinical indication for sampling or a chronic response to the presence of the shunt.

**Figure 4.**
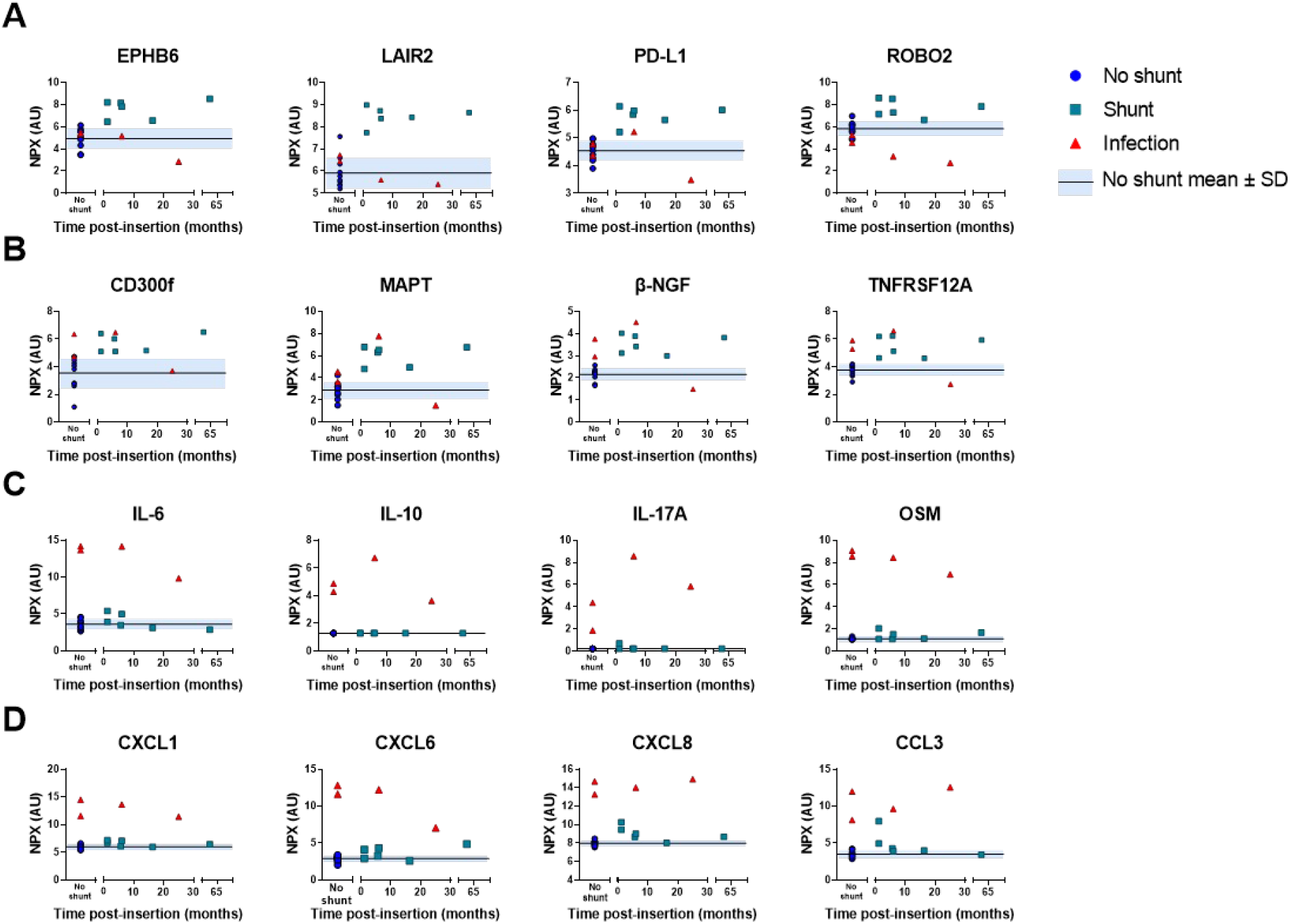
Representative proteins increased in CSF of individuals with shunts and/or brain infection over time. (A) Proteins increased only in shunted individuals without infection (B) Proteins increased in shunted and infected individuals (C) Cytokines increased in individuals with infection (D) Chemokines increased in individuals with infection

## Discussion

In this study, we investigated a panel of 182 immune and neurological markers in the CSF of shunted and unshunted patients. We show that the presence of shunt catheters within the cerebrospinal fluid gives rise to a neural response indicative of wound healing and cell migration. Importantly, these findings are distinct from the inflammatory response to infection and are independent of the time since the last surgery to either insert the shunt or revise it. If validated in larger clinical studies, these findings could facilitate further understanding of the biological mechanisms underpinning non-infected mechanical shunt obstruction. This is particularly pertinent since the frequency of implanted antibiotic-impregnated catheters has grown exponentially following a recent randomised-controlled trial where the incidence of shunt failure due to mechanical obstruction was noted to be higher in the antibiotic-impregnated shunts. The authors hypothesise that changes in CSF composition and flow due to low virulent pathogens restricted by biofilms in the shunt valve contributed. A comprehensive longitudinal study therefore, would shed light on the extent of the neuroinflammatory response as the catalyst for consequent shunt failure.

One of the strengths of this study is in its analysis of a wide panel of neural and immune biomarkers and in its timing of acquired CSF samples. The sampling of CSF from our non-shunted patients (n=10) was driven by therapeutic need i.e. to relieve the patient of their headaches experienced as a result of underlying idiopathic intracranial hypertension. This is representative of real-life clinical decision making. 3 patients in this group went on to have VP shunts. In the shunted patients, CSF sampling was taken at the time of clinical equipoise i.e. to rule in or out infection, and in non-infected patients, to aid in diagnosis of for e.g. shunt malfunction. This is indicative of real-world clinical difficulties and in our study, the utility of examining additional CSF biomarkers following initial CSF analysis was the pertinent question.

A strength of the study ethically, though weakness methodologically, is that the CSF samples obtained were taken only from patients in which it was deemed clinically necessary; in this patient cohort this meant when control IIH patients without shunts were experiencing symptoms and required diagnosis, or in shunted patients when the shunt malfunction resulted in symptoms returning. This means that samples are not perfectly matched, and that the possibility that these markers could be reflecting the difference in severity of symptoms between shunted and unshunted individuals, while unlikely, cannot be ruled out. In an ideal study samples would be taken from patients prior to shunting, while the shunt was working, and on blockage, however for ethical and practical reasons this was not possible. Similarly, the samples have been taken from a small group sizes of 10 unshunted and 10 shunted patients. We appreciate that this means the study needs to be confirmed in a larger, more defined cohort. However, the consistency of the biological responses seen between patients and the reliably constant pathways in which the biomarkers are involved suggest that the data will be robust in larger groupings.

Accurately diagnosing neurological infection is challenging and we chose a pragmatic approach of either bacteria seen on gram stain or positive bacterial culture. In cases of negative gram stain or culture, patients were deemed infected if they presented with a clinical picture highly suggestive of infection, supported by positive serum markers and CSF pleocytosis with a subsequent response to antibiotic treatment.

There are three non-exclusive potential reasons for shunted individuals showing the observed changes in their CSF. The first is that there is an ongoing allergic response to the shunt material. Given that allergic responses to silicone are uncommon, it is unlikely it has occurred in all of the tested shunt patients. Further, allergic responses are immune mediated and have a well-defined Th2 cytokine profile, usually accompanied by local eosinophils ^15–18^. Given that no Th2 cytokines were increased in the shunted patients, this is an unlikely explanation.

The second potential reason is that the increase in biomarkers and symptoms are both responses to increased intracranial pressure. This explanation would assume that the increase in biomarkers in shunted patients is a reflection of greater pathology in those patients in comparison to those not yet shunted. Unfortunately, we were not able to address this with the current dataset. Ideally, it would be investigated by taking study samples longitudinally from the same patients prior to shunting, while the shunt was working, and on blockage or infection in order to reduce variation due to individual patients. However, for ethical and practical reasons this was not possible. While we cannot rule this explanation out, the biological roles of the biomarkers equally do not support it.

The third potential explanation is that the protein changes are a window into an ongoing adaptive response to the presence of the shunt catheters in the cerebrospinal fluid. The identities of the proteins found indicate increased cellular locomotion and differentiation in shunted patients. It could be hypothesised that very small movements of the shunts during everyday life disturb the local tissue enough to cause ongoing tissue remodelling, which would account for the observed changes. This explanation is the most consistent with our findings and with cellular findings by other researchers in the field.

Our data show that there is a mild response to the presence of a shunt, consisting of neurological rather than immunological proteins. The most prominent of these is the tau protein encoded by the microtubule-associated protein tau (*MAPT*) gene, which is highly expressed in neurons but also in astrocytes and oligodendrocytes. Tau’s most well-known aspects of being associated with Alzheimer’s disease ^19,20^ and a potential biomarker for brain injury ^21^ may be linked to its importance in cell migration and differentiation, as has been demonstrated in the peripheral glial Schwann cells ^22^. Ephrin type-B receptor 6 (EPHB6) and CD300f (CMRF35-like molecule-1; CLM-1), two proteins that were also increased in shunted patients, are involved in cellular migration, consistent with the colonisation of the shunt with astrocytes and microglia described in previous studies ^4^. Similarly, β nerve growth factor (β-NGF) has been shown to be of multifaceted importance and widely expressed in neurons. Most pertinently to the current study, it is highly expressed in wound healing ^23^, and astrocytes treated with β-NGF have enhanced migration, leading to rapid wound closure in an *in vitro* model ^24^. Hence, the presence of MAPT, β-NGF, EPHB6 and CD300f is consistent with ongoing cell movement as part of the adaptive response to the presence of the shunt catheter within the cerebrospinal fluid.

In addition to biomarkers of cell movement, there were markers increased in shunted patients which show involvement of interacting systems, such as leukocyte-associated immunoglobulin-like receptor 2 (LAIR-2) and programmed cell death 1 ligand 1 (PD-L1). LAIR-2 is an immunoglobulin-like receptor which interacts strongly with collagen ^25^ and proteins with collagen domains such as complement components ^26^. It appears to interfere with thrombus formation through inhibiting collagen-platelet adhesion ^27^ and may play a role in modulating dendritic cell activation by competing with the DC-surface receptor LAIR-1 ^28^, while PD-L1 is primarily recognised as a dampener of T cell responses. It is possible that its expression is driven by the damage to astrocytes that occurs in patients during insertion and movement of implanted shunts ^29^.

In contrast to those patients with non-infected shunts, infection with a known bacterium, such as the cases of coagulase-negative staphylococci described here, gives rise to a classic inflammatory immune response as has been reported in a number of other studies in both humans and rodents ^13,30,31^. Notably, it is usually associated with a strong IL-6 and CXCL8 response (Cuff 2020).

As well as free-living bacteria, shunts can become colonised by bacteria that form biofilms on the silicone surface, and this can present a significant challenge when diagnosing neurological infection. Conventional symptoms such as fever, nuchal rigidity and photophobia are unreliable and microbiological culture may yield false negatives. In biofilms the bacteria are encased in a matrix including extracellular DNA and proteins which together function to protect individual cells from environmental insults including the immune system. They also express different genes compared to free-living bacteria ^8–10^, and can give rise to a distinct immune response, compared to the response to free living bacteria ^32^. Biofilms are associated with a distinctive innate immune response in animal models and patients. The immune response appears to include monocyte-derived suppressor cells (MDSCs) which secrete IL-10, ^13,36–38^. Given that we did not observe any increase in IL-10 in our patients it is unlikely that biofilms were present on the shunt catheters.

It was found previously that shunt placement can cause local scarring typified by an increase in glial cells in the local area ^39,40^. Increases in astroglia have also been reported to be a part of the IIH pathology ^41^ although the patient cohort in that study was predominantly shunted (15 of 18 patients) and so it is not possible to determine whether the results are due to IIH or the presence of shunts. These studies predominantly examined cellular changes and of necessity were performed in situations in which brain tissue could be taken, or in animal models. Our current study adds to the literature by exploring the new, complementary territory of soluble biomarkers and expanding the observations to include individuals without as severe a pathology. Importantly, our data rule out the hypothesis that ongoing responses to shunts are due to the presence of small amounts of bacterial proteins and lipids on the surface of the shunts stimulating a low-level inflammatory response, instead pointing to a long-lasting, non-inflammatory response of the local cells to the presence of the shunt.

## Data Availability

All data produced in the present study are available upon reasonable request to the authors.

## Data availablity

The datasets analysed during the current study are available from the corresponding author on reasonable request.

## Authors’ contributions

SC analysed and interpreted data and wrote the manuscript draft. JM collected the patient samples and curated patient medical data. ME provided scientific oversight. WG provided clinical oversight. All authors read, edited and approved the final manuscript.

## Acknowledgements

Dr Sam Loveless of the Welsh Neuroscience Research Tissue Bank was invaluable in curating the patient samples for this study.

## Supplementary figures

**Supplementary Table 1.**
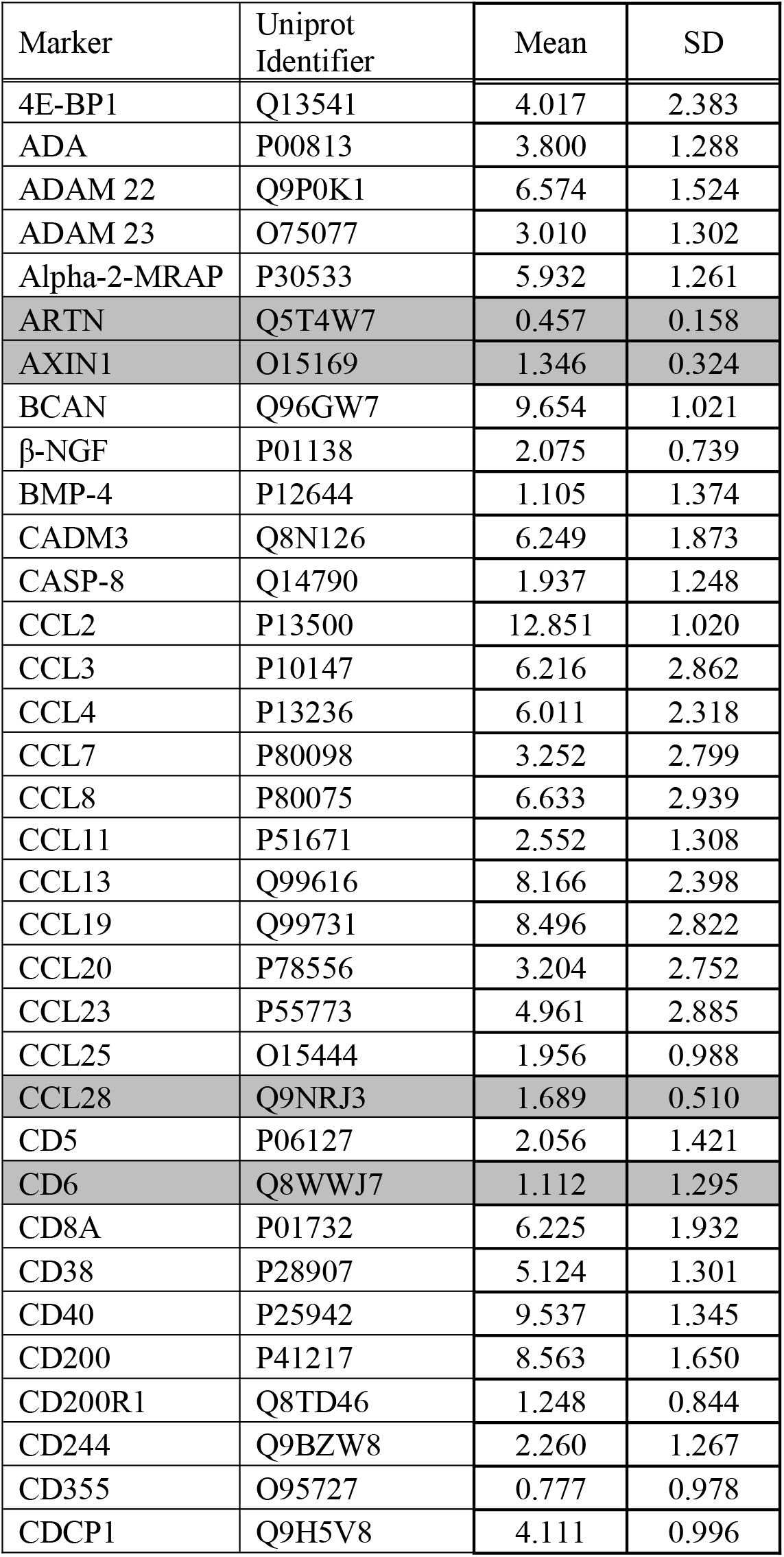
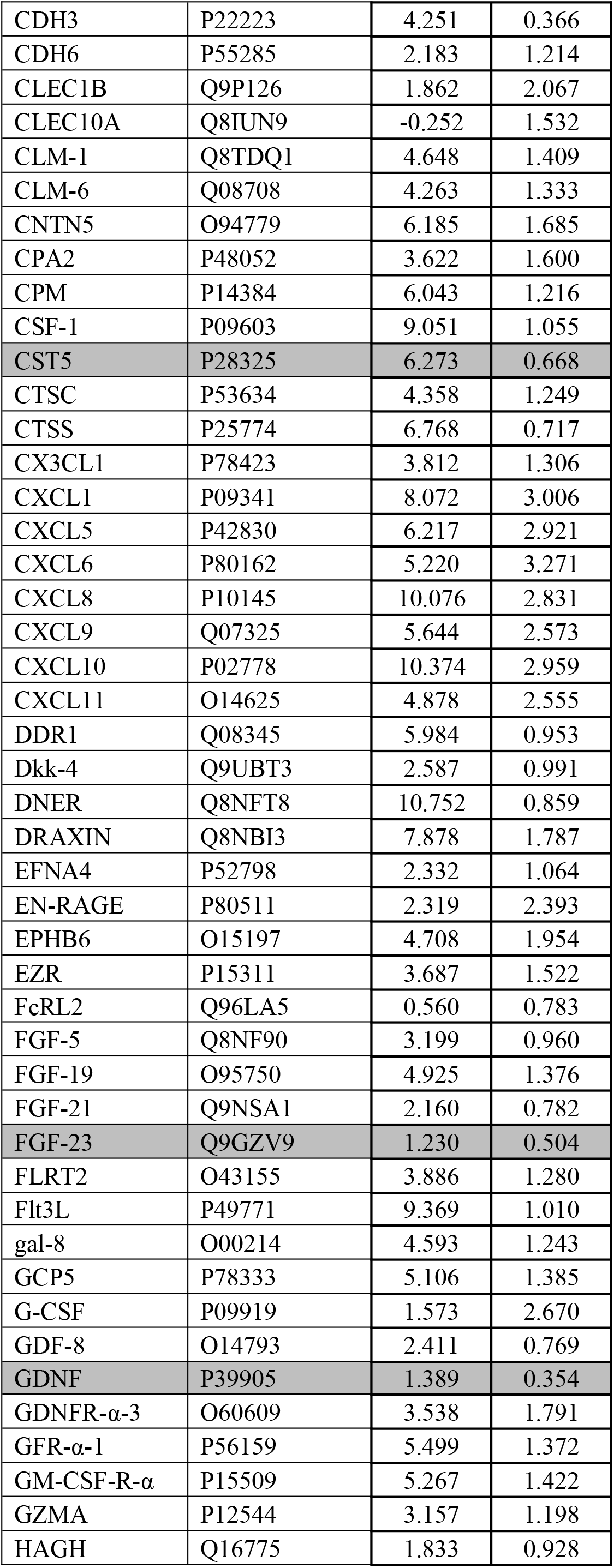
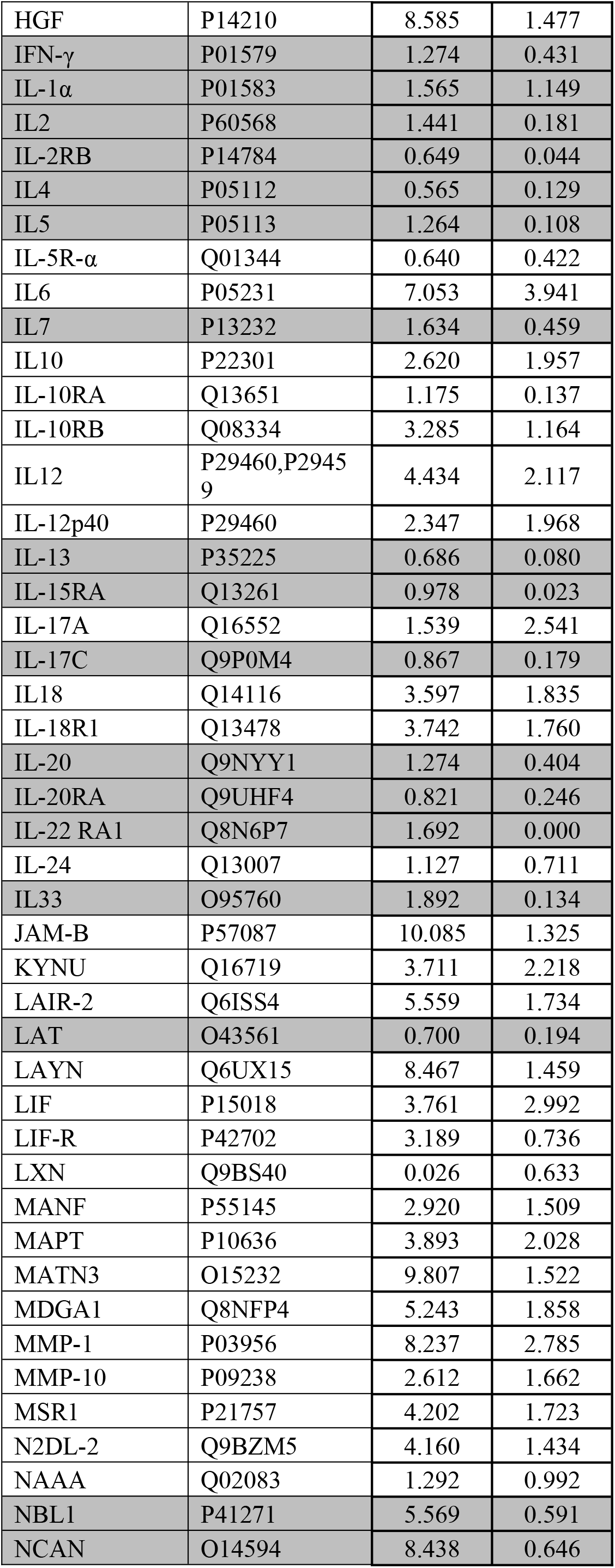
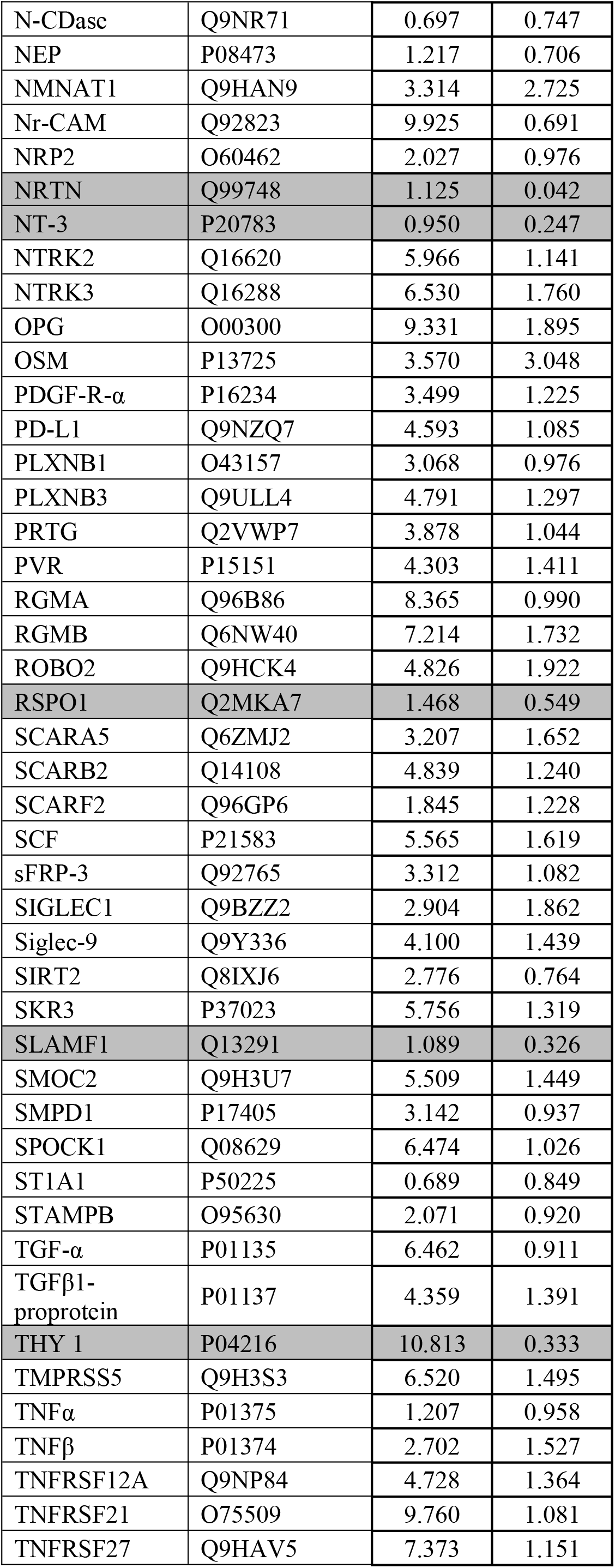
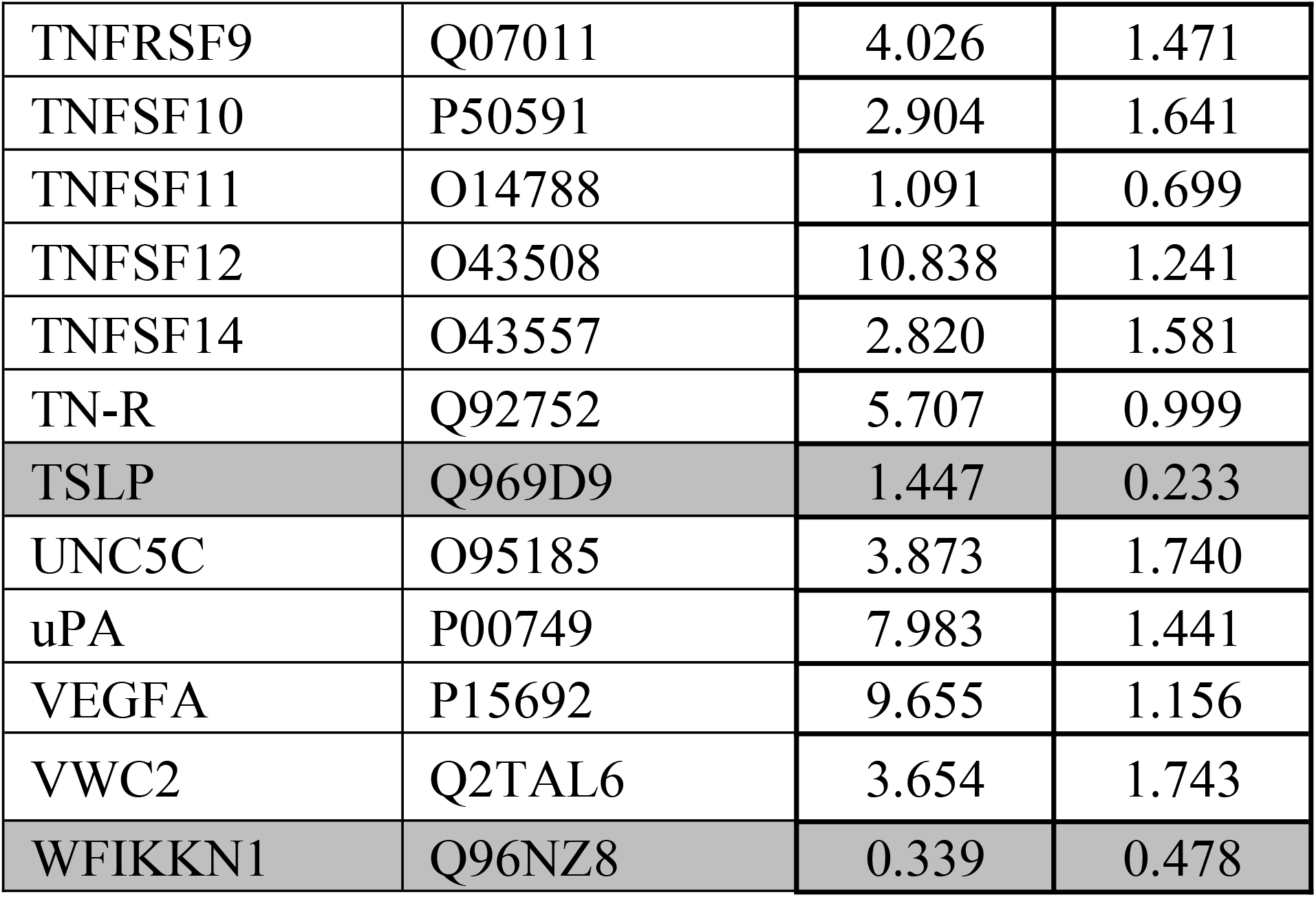
List of variables tested, with mean and SD. Concentrations are measured in normalised protein expression units. Those markers subsequently removed from the analysis are shaded in dark grey.

